# Using a Quality Improvement Approach to improve reporting of malaria deaths in Namutumba District, Eastern Uganda, 2022–2023

**DOI:** 10.1101/2024.05.21.24307657

**Authors:** Marie Gorreti Zalwango, Richard Migisha, Benon Kwesiga, Lilian Bulage, Daniel Kadobera, Alex Riolexus Ario

**Affiliations:** Uganda Public Health Fellowship Program, Uganda National Institute of Public Health, Kampala, Uganda

**Keywords:** Malaria deaths, Malaria, Quality improvement, Reporting, Namutumba, Uganda

## Abstract

**Background:** Reporting malaria deaths is critical for assessing prevention and case management interventions. In Uganda, malaria mortality is recorded in inpatient registers and reported through weekly and monthly surveillance reports. During a data quality assessment in Namutumba District in October 2022, we found more malaria deaths in health facility registers than were reported. We conducted a continuous quality improvement initiative to improve the accuracy of reported malaria deaths in Namutumba District.

**Methods:** We purposively selected 2 high-level health centers (HC) in Namutumba District that reported malaria deaths during September 2021–October 2022. We formed quality improvement teams (QIT) comprising clinical and statistical staff at the HC. We held focus group discussions with QITs to identify challenges with reporting malaria deaths, prioritized areas for improvement, and conducted root cause analysis. Using the plan, do, study, act (PDSA) cycle, we identified change ideas to address root causes.

**Interventions:** Challenges included knowledge gaps on malaria death definitions, lack of consequences for failing to document deaths, and unclear guidance on how to document deaths. Sustainable interventions identified included continuous medical education on malaria death definition, one-on-one mentorship of staff on documentation in inpatient registers, and weekly verification of inpatient register data, all implemented during November 2022–February 2023.

**Results:** Of the 36 malaria deaths that occurred during the baseline period (September 2021– October 2022), 25 (69%) were included in the weekly report, and four (11%) in the monthly report. Following the intervention implementation, all 7 malaria deaths recorded at the 2 health facilities during November 2022–February 2023 were reported in the weekly and monthly reports.

**Conclusion:** Continuous medical education, supervision and mentoring of HC staff, and clear and comprehensive guidance on documenting malaria deaths could facilitate improved malaria death reporting from HC in Uganda, enabling more accurate resource allocation for malaria control.

## Introduction

Malaria is one of the leading causes of mortality globally, responsible for 608,000 deaths in 2022 The vast majority (96%) of the reported deaths occur in 29 sub-Saharan Africa countries Uganda inclusive. Uganda ranks eighth globally among countries with the highest burden of malaria deaths. It causes 20-30 deaths weekly and around 1,300 annually, as per the Uganda Health Management Information System (HMIS) [1]. Due to challenges in reporting malaria deaths, the HMIS-reported numbers have been adjusted for underreporting, resulting in an estimated 19,600 deaths annually, according to the World Malaria Report [2].

Based on findings from a community study conducted in Namutumba District, several community malaria deaths were documented at health facilities but not recorded in the health information system due to deaths occurring before reaching high-level facilities [3]. Subsequently, we conducted a data quality assessment (DQA) from September 2021 to October 2022 with the aim of determining the accuracy of health facility malaria deaths data. By comparing registers and client cards to surveillance reports (HMIS 033b and HMIS 108) through the District Health Information System version 2 (DHIS2), the DQA revealed a discrepancy. Out of 36 malaria deaths recorded, 25 (69%) were included in the weekly report, and 4 (11%) in the monthly report, indicating an inconsistency in reporting between registers and surveillance reports.

The low reporting of actual numbers of malaria as the cause of deaths due to various challenges contribute to underestimation of the true magnitude of malaria deaths. Without accurate reporting of malaria mortality data at the district and national levels, planning for effective interventions to prevent malaria deaths remains insufficient, posing a challenge to the goal of malaria eradication by 2030. This initiative focused on improving the accuracy of reported malaria deaths in selected health facilities in Namutumba District during September 2021– October 2022, to provide a case study for improvement in other health facilities and districts in Uganda.

## Methods

### Project implementation setting

We conducted the quality improvement project in Namutumba District based on the previous low number of malaria deaths reported as per findings from the DQA. Namutumba District is located in the eastern part of Uganda, a region highly endemic for malaria [4, 5]. The district is made up of 2 constituencies and 20 sub-counties and has a total population of approximately 320,000 people. The population is served by 2 private hospitals, 1 public health center (HC) IV, seven HC IIIs, and 25 HC IIs [6]. However, due to the low economic status of the residents in the district, majority of the patients utilize services at the HC IV which serves as the district referral health unit beyond which patients are referred to higher facilities outside the district for further management. We conducted the project in two high-level HCs in Namutumba District because of their high capacity for severe malaria admissions and the high likelihood for malaria deaths. These were Namutumba HC III and Nsinze HC IV.

### Project implementation design

The quality improvement model used for our project focuses on 3 fundamental questions. What are we trying to accomplish? How will we know if a change is an improvement? and what changes can we make that will result in an improvement? Building on this information, we rejuvenated the Quality Improvement Team(QIT) for the selected facilities. This step was followed by collection of baseline data and we supported the teams to identify their problems and define their improvement objective. We further identified measures and changes to be made for improvement and implemented them using the Plan Do Study Act (PDSA) model.

### Continuous quality improvement team formation

We reconstituted the QIT at each facility to coordinate the implementation of project activities. The teams constituted included the malaria focal person, in-charge, health facility QI focal person, records assistant, and two clinicians at the outpatient clinic and inpatient ward. The roles of the QIT were as follows: 1) Team lead – responsible for assigning roles and adding a voice where it was required. Members on the project were asked to appoint their own leader and change when need arise. 2) Mentor – responsible for guiding the team in carrying out certain roles, and provided mentorship to the team members. 3) Data person – responsible for data collection and analysis of the collected data. 4) Support personnel – provided information or any additional assistance required to conduct the project.

### Baseline data collection

We analyzed health facility data for Nsinze HC IV and Namutumba HC III for the period of September 2021 to October 2022 to serve as our baseline for malaria deaths reporting. This involved review of the HMIS data for the HMIS 033b weekly surveillance report and the HMIS 105 monthly report, review of client cards for all admitted patients, review of the inpatient register and the outpatient register for cases that might have died before admission. This information helped the team to identify gaps in data capture and reporting. Using a score of 0-5 on the spider graph (5 being the highest ranking), the identified gaps were prioritized for problem analysis.

### Process mapping and gap identification

We reviewed the process of data capture in the primary data tools, data collection and data entry into the electronic HMIS system through focus group discussions with the facility teams. This was done to identify the step in the process with the most challenges affecting reporting for malaria deaths. Using the spider graph, gaps under each step of the process were charted for prioritization of the step with the most gaps.

### Problem analysis and root cause identification

Following prioritization of the problem, the QIT did problem analysis for the identified gaps to identify the root causes for low reporting for malaria deaths using the ‘five whys’ technique. For each of the problems, the team exhausted the possible causes until when the root cause was reached. These findings were displayed using a fish bone diagram.

### Goal refinement and change idea identification

Following problem identification, the QIT refined the goal for the project which was a clear statement of the intended improvement and how it would be measured. The goal had the following attributes: specific, measurable, achievable, realistic and time bound. The QIT proposed changes/interventions to the identified problems. Teams identified the different change ideas that would bring about the desired change. These change ideas were organized into groups, each of which represents a similar notion or approach to change, or change concept. We then ranked the proposed changes.

### Intervention tools

Based on the proposed changes, we identified tools required to implement the suggested changes. We focused on tools that were cheap and readily available to ensure sustainability of the project.

### Intervention monitoring and evaluation

We used the PDSA cycles to plan and test the identified interventions. Through the weekly and monthly data collection, analysis and interpretation, the team was able to assess each implemented intervention for a positive change. The plan was to maintain interventions that improved reporting for malaria deaths, while interventions that did not cause change were dropped. The final evaluation was done in March 2023 to ascertain whether there was an improvement in the reporting rates of malaria deaths.

### Ethical considerations

We obtained administrative clearance from Namutumba District Health Office and the health facility in charges for the selected health facilities. We further obtained verbal consent from the health facility staff that participated in the focus group discussions to generate existing gaps and required change ideas for improvement. Additional data was obtained from routinely collected malaria surveillance data in the health facility registers and the DHIS2 that is publicly available for analysis and use to inform public health interventions. The data was aggregated with no individual identifiers. This activity was reviewed by US Centers for Disease Control and Prevention (CDC) and was conducted consistent with applicable federal law and CDC policy.§ §See e.g., 45 C.F.R. part 46, 21 C.F.R. part 56; 42 U.S.C. §241(d); 5 U.S.C. §552a; 44 U.S.C. §3501 et seq. This determination was made because the project aimed to address a public health problem and had the primary intent of public health practice.

## RESULTS

### Baseline findings

During the baseline period reviewed, Namutumba HC III recorded 4 malaria deaths. Only 50% (2/4) of the confirmed malaria deaths were reported in the monthly HMIS 108 report while 100% (4/4) were reported in the weekly surveillance report. For Nsinze HC IV, 32 malaria deaths were recorded in the period of September 2021-October 2022. Findings from the baseline data collection revealed that only 19% (6/32) confirmed malaria deaths were reported as in the monthly surveillance report while 81% (26/32) were reported in the weekly surveillance reports. We noted that HMIS 033b was collected from both out-patient and in-patient registers while the HMIS 108 report was collected from only the in-patient register for both health facilities as opposed to utilization of the inpatient register as the official data source for malaria mortality.

### Process mapping and gap identification

Following review of the reporting system for malaria deaths, gaps were identified at the steps of data entry in the primary registers and data compilation from the registers. Other steps like data entry on hard copy reports and entry of data in the electronic HMIS had no gaps identified (Figure 1). This informed our choice to focus on data entry into the inpatient register for root cause analysis since this is the primary source document for malaria deaths data.

**Figure 1:**
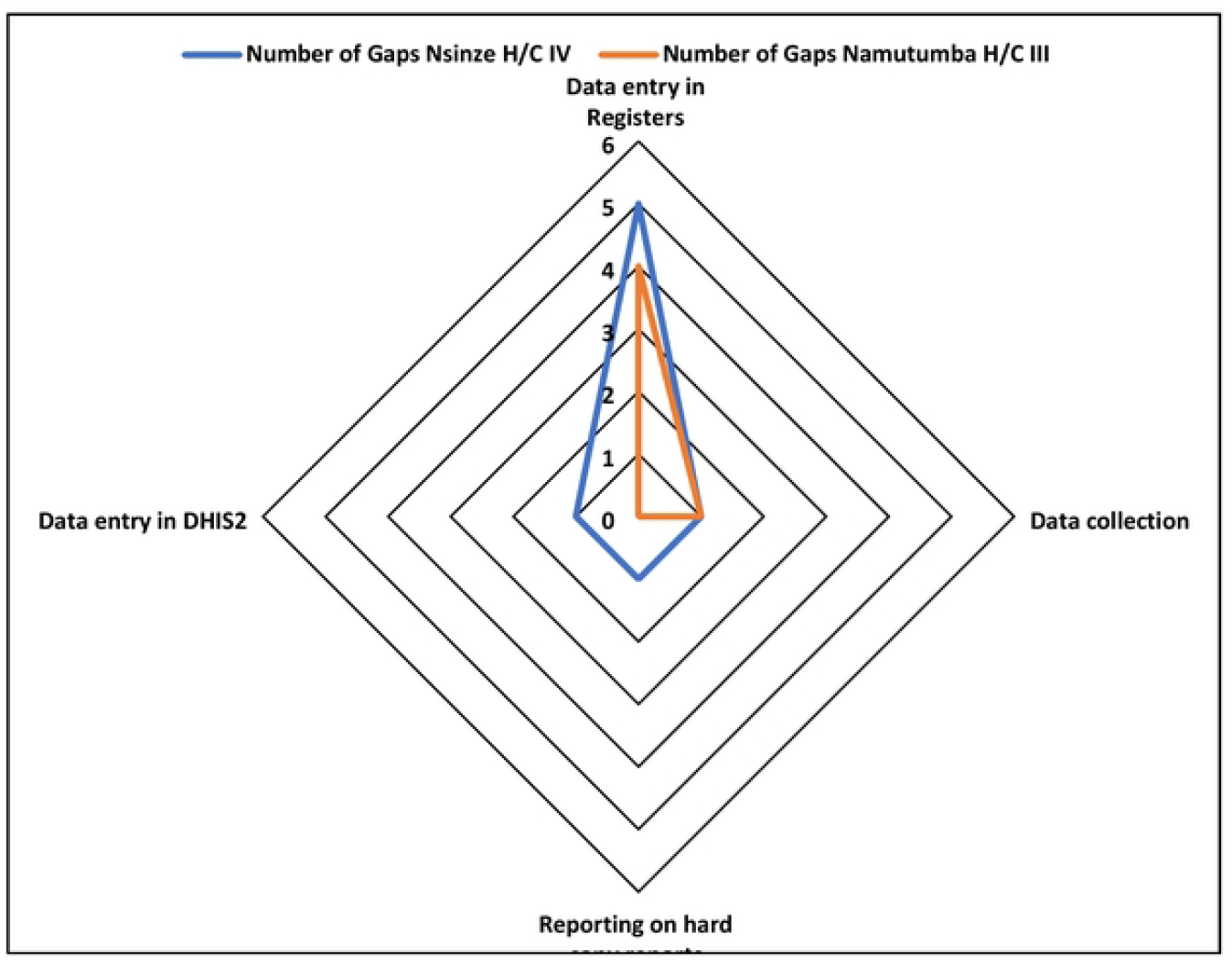
Number of gaps identified during the process mapping activity, Namutumba District, Eastern Uganda, September 2021-October 2022.

### Root causes affecting entry of malaria deaths data in the in-patient registers for Namutumba District, September 2021–October 2022

As depicted in figure 2 below, the root cause of failure to enter malaria death data included:

**Figure 2:**
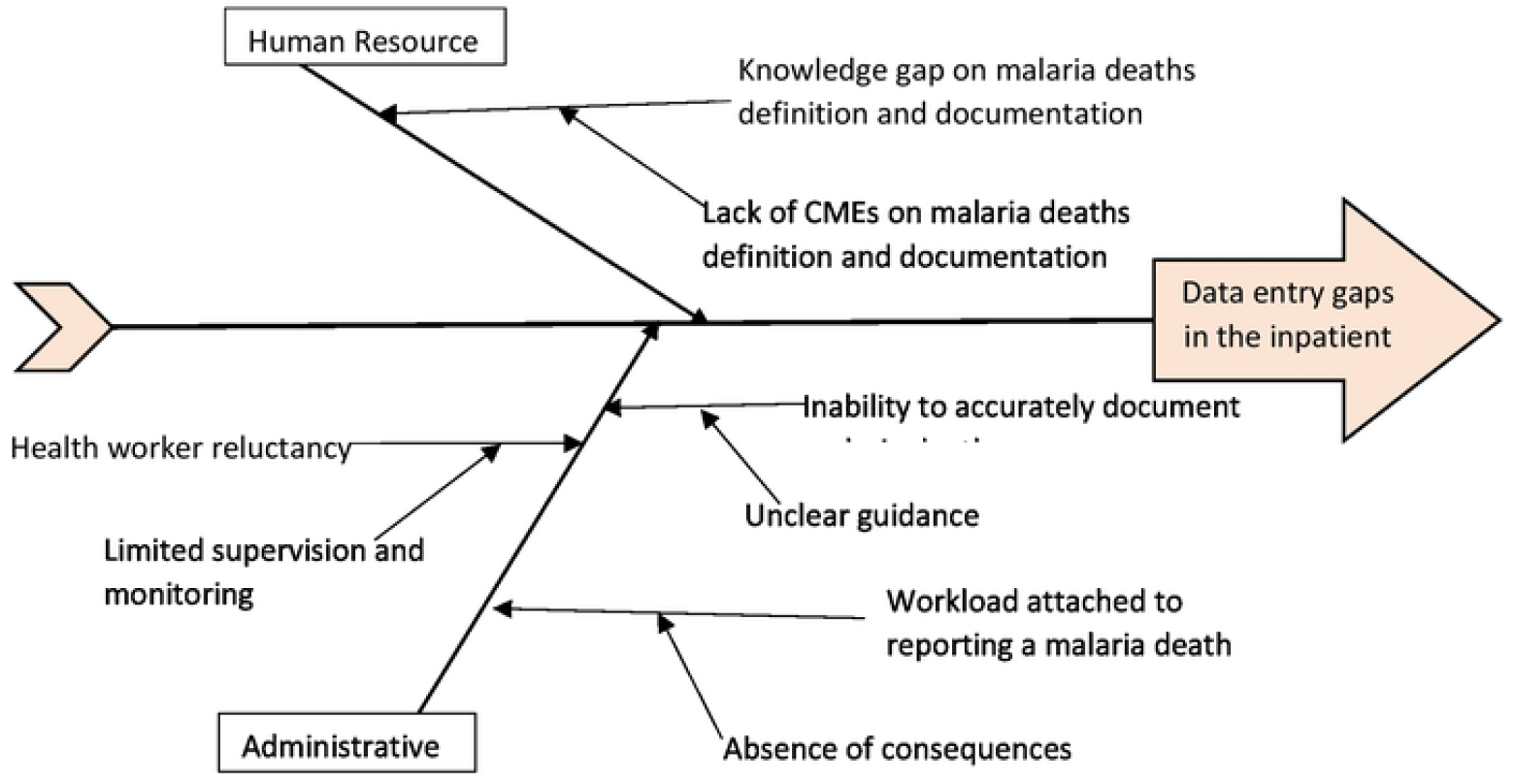
Root causes for the gaps identified during the problem identification at the health facilities in Namutumba District, Eastern Uganda, September 2021–October 2022.

**Figure 3:**
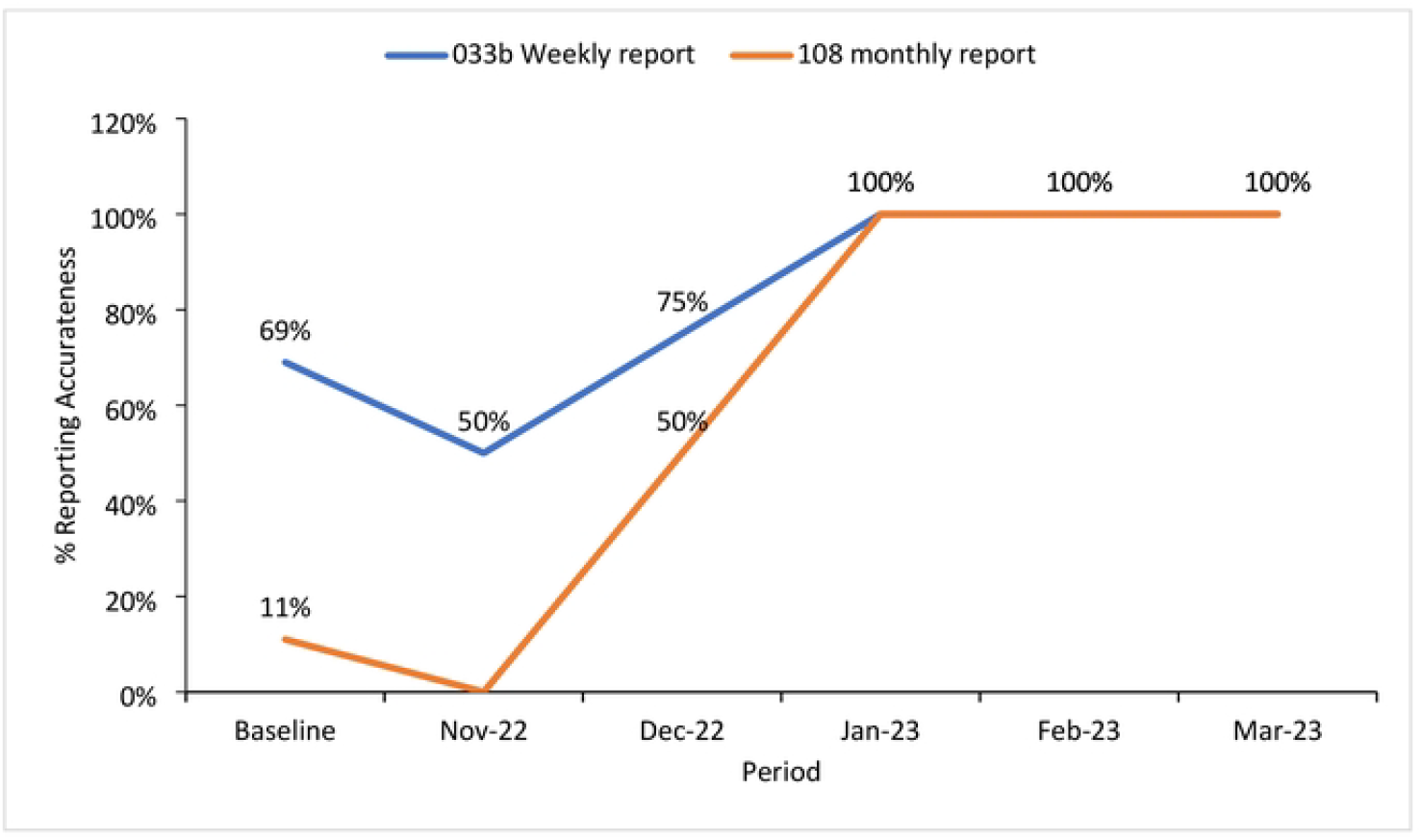
Changes in malaria mortality reporting for the health facilities in Namutumba District, Eastern Uganda, November 2022–March 2023.

#### Lack of continuous medical education on malaria deaths definition and documentation

Insufficient continuous medical education (CMEs) on the definition and documentation of malaria deaths led to health workers documenting complications rather than malaria as the cause of death in primary registers, despite malaria being the primary diagnosis. The knowledge gap resulting from the lack of CMEs contributed to low documentation and reporting of malaria deaths.

#### Absence of consequences for non-reporting of malaria deaths

The absence of consequences for non-reporting of malaria deaths led to reluctance among health workers to document such deaths, as it involved additional paperwork, including filling out medical certificates of cause of death and entering data into the DHIS2.

#### Limited supervision and monitoring performance

Limited supervision and monitoring performance of health workers contributed to staff reluctance in accurately documenting and reporting malaria deaths.

#### Unclear guidance on malaria death documentation

Severe malaria deaths are expected to be recorded in the in-patient register; however, deaths occurring at health facility gates, outpatient department, and on the facility premises after referral were often not documented or recorded in the outpatient register. This inconsistency resulted in discrepancies between weekly and monthly reports, as the monthly report relies on both inpatient and outpatient registers. Similar challenges arose for deaths occurring on the way to the health facility or at home, as there was no clear guidance on documenting such deaths happening outside the inpatient wards and in the community.

Following the brainstorming exercise, a range of change interventions were identified for improving reporting of malaria death data. These included:

a. Conducting regular CMEs on malaria death definition, recording, and reporting
b. Conducting CMEs to guide on the documentation of death on arrival and after referral
c. Conducting one on one mentorship on malaria deaths documentation
d. Establishment of focal persons to ensure timely update of malaria deaths in the IPD register
e. Conducting weekly malaria death data reviews
f. Strengthening the system of support supervision and monitoring

The change interventions were then turned into activities with clear indicators and means of verification and responsible persons identified for each activity (Table 1)

**Table 1.** Quality improvement interventions and indicators.

| **Intervention** | **Activity Description** | **Responsible Person** | **Indicator** | **Means of verification** |
| --- | --- | --- | --- | --- |
| Conducting regular CMEs on malaria death definition, recording, and reporting | Bi-monthly CMEs on malaria deaths definition, documentation and reporting. Using a CME schedule, notification was made to all health facility team by in charges to ensure attendance. | Clinician responsible for CMEs | Proportion of health facility staff with knowledge of malaria death definition, recording, and reporting | Inpatient register |
| Conducting CMEs to guide on the documentation of death on arrival and after referral | The QIT discussed and agreed on the best way to document malaria death on arrival and after referral. Since there was no clear guidance, the team agreed that malaria deaths on arrival with positive malaria results and those dying in the resuscitation room would be documented in a special way in the inpatient register clearly indicating the place of deaths in the comment section. Patients dying on the health facility premises following referral would have their outcome changed from referred to Died and captured as a health facility death. These resolutions were provided to the rest of the team during the bi-weekly CMEs to ensure implementation by every staff member. | Clinician responsible for CMEs | Proportion of malaria deaths at the facility documented in the inpatient register. | Inpatient register |
| Conducting one on one mentorship on malaria deaths documentation | Mentorship of clinicians (medical officers, nurses and midwives) and medical records staff on required data parameters for malaria deaths documentation | Clinician responsible for CMEs | % of health workers with competency to document malaria deaths completely and accurately | Inpatient register |
| Establishment of focal persons to ensure timely update of malaria deaths in the IPD register | Assigned focal persons reviewed the registers after recording a death to ensure completeness and accurateness of documentation | Malaria Focal Person | Proportion of malaria deaths at the facility documented completely and accurately in the inpatient register. | Inpatient register |
| Conducting weekly malaria death data reviews | Weekly review of inpatient client cards and Outpatient registers to ensure that all malaria deaths were documented in the inpatient register. | Quality Improvement Team | Proportion of malaria deaths at the facility documented in the inpatient register. | Inpatient register |
| Strengthening the system of support supervision and monitoring | Weekly review of the inpatient registers to ensure that all malaria deaths recorded have medical certificates of cause of death filled and filed | Health Facility In charges | % malaria deaths reported in HMIS033b | HMIS 033b report<br>Inpatient register |

### Final evaluation of the quality improvement project

Following implementation of QI project in Namutumba District, there was an improvement in the reporting of malaria deaths from 69% for the weekly report and 11% for the monthly report to 100% during the period of November 2022 to March 2023 with all the seven malaria deaths recorded and reported in the weekly and monthly reports. All the seven malaria deaths that happened in the health facilities were appropriately documented in the primary source document (inpatient register) and reported in the weekly surveillance and monthly reports for all the 5 months of project implementation.

## Discussion

The project identified gaps and challenges in reporting malaria deaths, leading to low rates in both weekly and monthly surveillance reports. Contributing factors included the absence of Continuous Medical Education (CME) on malaria deaths, limited supervision, lack of consequences for non-reporting, and unclear guidance on documenting deaths in specific scenarios. After implementing proposed changes, there was an improvement in reporting malaria deaths, enhancing data quality at health facility, district, and national levels. This improvement could positively impact planning efforts by various stakeholders.

The study revealed a knowledge gap among health workers regarding the definition of malaria death due to the lack of CME on this topic. This knowledge gap resulted in misclassification and the subsequent underestimation of the burden of malaria deaths. According to the district health information system version 2 (DHIS2), a malaria death is defined as any death with a positive malaria test in a reporting health facility or community [7, 8]. Severe malaria that causes deaths presents with numerous complications like anaemia, respiratory failure, impaired consciousness and renal failure [9]. This complexity contributes to potential misclassification, particularly in the presence of a knowledge gap on the case definition of malaria death. Similar challenges have been observed in India, where a study highlighted the difficulty in defining and recording malaria deaths, because individuals with malaria may also suffer from other illnesses concurrently or in quick succession [10]. Regular CMEs are essential to ensure that all attending clinicians comprehend the accurate case definition for malaria deaths, facilitating accurate documentation and reporting.

Our study highlighted the lack of clear guidance on the documentation of malaria deaths upon arrival at the facility and deaths at the health facility premises after referral. As data on malaria deaths is extracted from the inpatient register (HMIS 075) and reported in the HMIS 108 report (Inpatient report), cases that do not reach the inpatient unit are often missed. Deaths on arrival might be recorded in the outpatient registers and included in the weekly surveillance report from the outpatient register, leading to inconsistencies between the HMIS 033b and HMIS 108 reports. The US Presidential Malaria Initiative (PMI), Roll Back Malaria (RBM) and Measure Evaluation have provided guidance for Evaluating the Impact of National Malaria Control Programs in Highly Endemic Countries. They highlight challenges in malaria-specific mortality surveillance and recommend using multiple data sources to compliment limitation from each method. Recommended sources in addition to the health information system include use of verbal autopsies, health and demographic surveillance systems (HDSS), civil registration and vital statistics systems and lastly the use of all child mortality as an impact indicator [11].

The study further revealed unclear guidance on documentation and recording of community malaria deaths. According to the NMCD, community/village health teams (VHTs) should follow up cases at home to ensure treatment compliance and recovery and to report any deaths due to malaria at home[7]. VHTs are required to report their data to health facilities quarterly; however, very few of them do report citing numerous challenges, including lack of transport, since in Uganda they are volunteers with no pay and with little supervision[12-14]. Scholars from other countries have suggested monetary facilitation and supervision to ensure they execute their roles[12, 15]. Moreover, the quarterly reporting system lacks timeliness for informing interventions. As such, exploring more frequent reporting mechanisms could enhance the capture of accurate and up-to-date information. Establishing clear guidance on the optimal reporting of community malaria deaths and issuing an official memo to all districts on this matter could ensure that reported malaria death numbers accurately reflect the burden of malaria deaths, and facilitating effective planning.

Our study revealed staff reluctance to document malaria deaths due to the additional workload involved in reporting, such as completing the medical certificate of cause of death. In a study to evaluate the impact of system failures on health workers, researchers noted that a failure in the complex system health workers work from causes physical and emotional burnout which may manifests as decreased production and lack of motivation. A system approach to existing challenges affecting health workers was proposed for improved performance[16]. Similarly, in our setting, addressing workload challenges by encouraging teamwork coupled with enhanced supervision and mentorship, could improve health worker productivity and ensure accurate reporting for effective planning of required interventions

### Study limitations

Our study faced limitations regarding the timely reporting of community malaria deaths due to a lack of guidance from the Ministry of Health on capturing these data at health facilities on a weekly and monthly basis. This limitation could have led to an underestimation of malaria deaths in Namutumba District. However, we minimized this limitation by strengthening the documentation of all malaria deaths occurring on arrival and immediately after referral from the health facility.

## Conclusion

There was improvement in malaria mortality reporting attributed to continuous medical education, supervision and monitoring performance of health facility staff, clear and comprehensive guidance on documentation of malaria deaths and weekly data reviews. Ensuring regular CMEs, data review, support supervision, and comprehensive guidance on documenting malaria deaths could enhance reporting, aiding planning, and reinforcing efforts to achieve the malaria eradication target by 2030.

## Data Availability

The datasets upon which our findings are based belong to the Uganda Public Health Fellowship Program. For confidentiality reasons the datasets are not publicly available. However, the data sets can be availed upon reasonable request from the corresponding author and with permission from the Uganda Public Health Fellowship Program.

## List of Abbreviations

CQI: Continuous Quality Improvement
DHI: Division for Health Information
DHIS2: District Health Information System 2
DHT: District Health Team
HMIS: Health Management Information System
IPD: In-patient Department
MOH: Ministry of Health
NMCD: National Malaria Control Division
OPD: Out-patient Department
PDSA: Plan Do Study Act
VHT: Village Health Team

## Declarations

### Consent for publication

Not applicable.

### Funding and Disclaimer

This project was supported by the United States Agency for International Development (USAID)/President’s Malaria Initiative (PMI) through the US Centers for Disease Control and Prevention Cooperative Agreement number GH001353 through Makerere University School of Public Health. Its contents are solely the responsibility of the authors and do not necessarily represent the official views of the US Centers for Disease Control and Prevention, the Department of Health and Human Services, Makerere University School of Public Health, or the MoH. The staff of the funding body provided technical guidance in the design of the study, ethical clearance and collection, analysis, and interpretation of data, and in writing the manuscript.

### Authors contribution

GMZ collected data under technical guidance and supervision of RM, BK, DK, LB and ARA. GMZ analyzed and interpreted the data. GMZ drafted the manuscript. GMZ, RM, LB, ARA, critically reviewed the manuscript for intellectual content. All co-authors read and approved the final manuscript. GMZ is the guarantor of the paper.

## Acknowledgements

We appreciate the Nsinze HC IV and Namutumba HC III health facility staff for their active participation and implementation of the project. More appreciation to the Namutumba District health Office for their leadership and guidance most especially the District Malaria Focal Person.

